# Delirium Incidence, Duration and Severity in Critically Ill Patients with COVID-19

**DOI:** 10.1101/2020.05.31.20118679

**Authors:** Sikandar H Khan, Heidi Lindroth, Anthony J. Perkins, Yasser Jamil, Sophia Wang, Scott Roberts, Mark Farber, Omar Rahman, Sujuan Gao, Edward R. Marcantonio, Malaz Boustani, Roberto Machado, Babar A. Khan

## Abstract

**Background:** Delirium incidence, duration and severity in patients admitted to the intensive care unit (ICU) due to COVID-19 is not known.

**Methods:** We conducted an observational study at two large urban academic Level 1 trauma centers. Consecutive patients admitted to the ICU with a positive SARS-CoV-2 nasopharyngeal swab polymerase chain reaction test from March 1^st^, 2020 to April 27, 2020 were included. Individuals younger than 18 years of age, without any documented delirium assessments (CAM-ICU), or without a discharge disposition were excluded. The primary outcomes were delirium rates and delirium duration and the secondary outcome was delirium severity. Outcomes were assessed for up to the first 14 days of ICU stay.

**Results:** Of 243 consecutive patients with confirmed COVID-19 admitted to the ICU, 144 met eligibility criteria and were included in the analysis. Delirium occurred in 73.6% (106/144) and delirium or coma occurred in 76.4% (110/144). Sixty-three percent of patients were positive for delirium on the first CAM-ICU assessment. The median duration of delirium and coma was 7 days (IQR: 3-10), and the median delirium duration was 5 days (IQR: 2-7). The median CAM-ICU-7 score was 6 (IQR: 4-7) representing severe delirium. Mechanical ventilation was associated with greater odds of developing delirium (OR: 42.1, 95%CI: 13.0-137.1). Mortality was 26.4% in patients with delirium compared to 15.8% in patients without delirium.

**Conclusions:** 73.6% of patients admitted to the ICU with COVID-19 experience delirium that persists for approximately 1 week. Invasive mechanical ventilation is significantly associated with odds of delirium. Clinical attention to prevent and manage delirium and reduce delirium duration and severity is urgently needed for patients with COVID-19.

## Introduction

The Severe Acute Respiratory Syndrome 2 (SARS-CoV-2) novel coronavirus (COVID-19) has emerged as a global pandemic and is associated with rapid spread, severe respiratory failure and significant morbidity and mortality.^1,2^ As clinical experience with COVID-19 grows, neurologic manifestations of the disease are receiving increased attention. A recently published small case series from France reported delirium occurred in 26/40 (65%) of patients admitted to the intensive care unit with COVID-19.^3^ However, the duration and severity of delirium in critically ill COVID-19 patients have not been well described.

Delirium is a serious neurologic syndrome independently associated with longer duration of mechanical ventilation, prolonged ICU and hospital stays, increased mortality, and institutionalization after discharge.^4–8^ Increasing levels of delirium severity and duration amplify these outcomes, and are independently associated with worsening cognitive and functional outcomes post discharge.^9–12^ Prior to COVID-19, the prevalence of delirium in mechanically ventilated patients has been decreasing from a historically high rate of 80% to a range of 16.5–33%.^13–19^ In the setting of the current global health crisis, hospital resources have been stretched to their limits to meet the needs of a large number of critically ill patients. The unintended impact of limited resources on clinical practice has raised concerns that current ICU delirium rates have returned to the historically high levels.^20–22^ As of May 23, 2020, there are 1.64 million confirmed COVID-19 cases in the US and approximately 12% of COVID-19 patients required intensive care unit (ICU) level care.^1,2,23^ In this context, delirium is likely to pose a long-term public health challenge if rates in the United States are as high as recently reported in France.

Therefore, we conducted this study at two large academic health systems in urban Midwest to measure incidence of delirium, delirium duration and delirium severity, and investigate risk factors associated with delirium in critically ill patients admitted with COVID-19.

## Methods

The observational study was conducted at two large, urban, academic, Level 1 trauma centers (Indiana University Health Methodist Hospital and Eskenazi health) serving as major referral hospitals for the state of Indiana and affiliated with Indiana University School of Medicine (Indianapolis, Indiana). Methodist Hospital is an 802-bed quaternary care referral center with an average of 150 ICU admissions per month. Eskenazi Health is a 336-bed safety net hospital with an average of 120 ICU admissions per month. The study received ethical approval from the Institutional Review Board at Indiana University. All consecutive patients admitted to the intensive care units of Methodist Hospital and Eskenazi Health with a positive result by SARS-CoV-2 nasopharyngeal swab polymerase chain reaction test from March 1, 2020 to April 27, 2020 were included. Exclusion criteria were: patients under the age of 18, admitted after April 27, 2020, patients with no delirium assessments recorded in the electronic medical record for the duration of the follow up period, and those still admitted to the ICU or hospital at the end of the study period. We excluded patients remaining admitted to the ICU or hospital to accurately identify delirium duration and to prevent confounding of downstream effects of delirium on mortality and length of stay. Clinical outcomes were followed up until April 29, 2020 (date inclusive) or until the patient transferred out of the ICU.

## Exposures and Outcomes

The main exposure variables were patients’ demographics, comorbidities, laboratory results and severity of illness at admission. The primary outcomes were rate of delirium and delirium/coma duration during the first 14 days of admission to the ICU. Delirium/coma duration was defined by the number of days the patient was alive and had documented delirium or coma, representing duration of abnormal cognitive status. Patients who were discharged from the intensive care unit prior to 14 days did not have subsequent delirium or coma assessments performed outside the ICU. Coma was assessed using the Richmond Agitation Sedation Scale (RASS) and delirium was identified through the Confusion Assessment Method for the ICU (CAM-ICU). Coma was defined as a RASS score of –4 or –5, making patients ineligible for a CAM-ICU screening, while patients with a RASS score of –3 or greater were eligible for a CAMICU assessment.^24,25^ The CAM-ICU score was determined by examining the patient for (a) acute or fluctuating changes in mental status, (b) inattention, (c) altered level of consciousness, and (d) disorganized thinking. Patients were considered delirious if they displayed (a) and (b), plus (c) and/or (d) on the CAM-ICU. ICU nurses administered the RASS and CAM-ICU twice daily (around 0700 to 0800, and then between 1900 to 2000) to measure level of consciousness and delirium, respectively. These standardized and validated screening tools were implemented in our healthcare systems in 2011 and are normally obtained throughout the ICU stay. Hyperactive delirium was defined as a RASS score of +1 to +4 at the time of positive CAM-ICU, and hypoactive delirium was defined as a RASS score –3 to 0 with a positive CAM-ICU score. The secondary outcome of delirium severity was assessed using the Confusion Assessment Method for the Intensive Care Unit-7 (CAM-ICU-7) which requires all components of the CAMICU to be assessed for each patient rather than a dichotomous CAM-ICU positive or negative result. The CAM-ICU-7 was implemented into the electronic medical record at Eskenazi Health in 2017, and is assessed twice daily in the subset of patients receiving care at this hospital site. CAM-ICU-7 scores range from 0 to 7, with 0–2 indicating no delirium, 3–5 mild to moderate delirium, and 6–7 as severe delirium.^12^

## Data Collection

Research assistants familiar with electronic medical systems at the hospitals (Cerner PowerChart, Epic Health Systems) abstracted study data from the medical record, including CAM-ICU assessments performed by clinical nurses, and results were entered directly into an electronic REDCap database. Data obtained from the medical record included patient demographics (age, gender, self-reported race), insurance status, comorbidities, vital signs, laboratory and imaging results (within 24 hours of ICU admission), level of consciousness (RASS), date/time and results of delirium assessments for up to the first 14 days of ICU stay (overall CAM-ICU positive or negative, including CAM-ICU features as applicable: altered mental status, disorganized thinking, altered level of consciousness, disorganized thinking; and CAM-ICU-7 scores), SARS-CoV-2 test results, and dates of admission and discharge from hospital and ICU. Date of death during the hospitalization was also recorded including level of care at time of death (ICU vs. non-ICU). Comorbidities are presented as Charlson Comorbidity Index using diagnoses lists documented in the medical record. Acute Physiology and Chronic Health Evaluation Score (APACHE-II) was calculated using laboratory values, vital signs, and neurologic assessments from first 24 hours of ICU admission.

## Statistical Analysis

Demographic and clinical characteristics were compared between patients who had delirium positive and those without delirium using two-sample *t*-tests (normal data) and Wilcoxon Rank Sum tests (skewed data) for continuous outcomes or Fisher’s Exact test for categorical variables. Summary statistics including median and inter-quartile range (IQR) were provided for patients with delirium. Logistic regression was used including demographic or clinical characteristics that were significantly different between patients with delirium and those without delirium as independent variables to identify factors associated with delirium.

## Results

Two-hundred forty-three consecutive patients with COVID-19 were admitted from March 1, 2020 to April 27, 2020 to the ICUs at two hospital systems. We excluded 99 patients; 21 did not have any delirium assessments, and 78 remained admitted at the end of the follow up period (see Supplementary Figure 1). In total, 144 patients comprised the study cohort. Demographics and clinical characteristics for the cohort are presented in Table 1. The mean age of the cohort was 58 years (SD = 15.8), 42.4% were female, 50% African American and 14.1% Hispanic, 32.6% utilized commercial insurance, and 21.5% Medicare. The median Charlson Comorbidity Index score was 1 (IQR: 0–2), with hypertension (59.7%), obesity (56.1%), tobacco use (27.1%), and chronic lung disease (26.4%) the most frequent comorbid conditions. The median APACHE-II score was 17 (IQR: 13–24), and 73% of patients in the cohort underwent invasive mechanical ventilation. Cerebrovascular accident (ischemic or hemorrhagic) was identified in 1.4% (2/144) of patients.

**Table 1.** Characteristics of Critically Ill Patients Admitted with COVID-19 (n = 144)

| Demographic/Clinical Characteristic | Total (n=144) | Delirium Positive (n=106) | Delirium Negative (n=38) |
| --- | --- | --- | --- |
| **Age, mean (SD)** | 58.0 (15.8) | 58.9 (15.5) | 55.7 (16.5) |
| **Age, stratified n (%)** |  |  |  |
| 18-49, n (%) | 44 (30.6) | 31 (29.2) | 13 (34.2) |
| 50-64, n (%) | 48 (33.3) | 32 (30.2) | 16 (42.1) |
| 65+, n (%) | 52 (36.1) | 43 (40.6) | 9 (23.7) |
| **Sex, n (%)** |  |  |  |
| Female | 61 (42.4) | 48 (45.3) | 13 (34.2) |
| **Race, n (%)** |  |  |  |
| African American | 71 (50.0) | 50 (48.1) | 21 (55.3) |
| Caucasian | 43 (30.3) | 32 (30.8) | 11 (28.9) |
| Hispanic | 20 (14.1) | 15 (14.4) | 5 (13.2) |
| **Insurance, n (%)** |  |  |  |
| Medicare | 31 (21.5) | 22 (20.8) | 9 (23.7) |
| Medicaid | 19 (13.2) | 15 (14.2) | 4 (10.5) |
| Medicare & Medicaid | 15 (10.4) | 11 (10.4) | 4 (10.5) |
| Commercial | 47 (32.6) | 33 (31.1) | 14 (36.8) |
| Self-pay | 17 (11.8) | 13 (12.3) | 4 (10.5) |
| **Comorbidities, n (%)** |  |  |  |
| Hypertension | 86 (59.7) | 66 (62.3) | 20 (52.6) |
| Diabetes | 58 (40.3) | 41 (38.7) | 17 (44.7) |
| Obesity, BMI >30 | 74 (56.1) | 59 (60.2) | 15 (44.1) |
| Tobacco use | 39 (27.1) | 27 (25.5) | 12 (31.6) |
| COPD or Asthma | 38 (26.4) | 29 (27.4) | 9 (23.7) |
| Chronic kidney disease | 22 (15.3) | 17 (16.0) | 5 (13.2) |
| Chronic Heart Failure | 18 (12.5) | 14 (13.2) | 4 (10.5) |
| Cardiac Artery Disease | 17 (11.8) | 15 (14.2) | 2 (5.3) |
| Dementia | 5 (3.5) | 4 (3.8) | 1 (2.6) |
| **Charlson Comorbid Index** | 1 (0-2) | 1 (0-2) | 1 (0-2) |
| **APACHE II \*** | 17 (13-24) | 19 (15-26) | 13.5 (9-17) |
| **Laboratory Values, Clinical and Respiratory Characteristics, median (IQR)** |  |  |  |
| White blood cell count x10<sup>9</sup>/L | 8.7 (6.5-12.6) | 8.8 (6.5-13.6) | 7.5 (6.5-9.9) |
| Glasgow Coma Scale (0-15) | 10.0 (7.0-15.0) | 10.0 (6.0-14.0) | 15.0 (14.0-15.0) |
| Partial Pressure of Oxygen (PaO<sub>2</sub>) mmHg | 73.0 (59.0-105.0) | 73.0 (61.0-106.0) | 74.5 (53.0-95.0) |
| PaO<sub>2</sub>:FiO<sub>2</sub> ratio* | 96.7 (73.8-150.0) | 90.0 (70.0-140.0) | 134.4 (95.0-188.9) |
| RASS (14 days) | -1, (-2, 0) | -1.6 (-2.5, -0.8) | 0 (0, 0) |
| Invasive mechanical ventilation* | 105 (72.9) | 97 (91.5) | 8 (21.1) |
| Presence of Shock | 21 (14.6) | 19 (18.1) | 2 (5.1) |

Table 1 describes the demographic and clinical characteristics of the overall cohort. Laboratory values, clinical, respiratory characteristics, and severity of illness measurements represent data from first 24-hours of ICU admission. Delirium status determined by a positive CAM-ICU clinical assessment. RASS scores were calculated using scores for up to 14 days. Severity of illness, Glasgow Coma Scale, RASS scores, percent mechanically ventilated, and PaO<sub>2</sub>:FiO<sub>2</sub> were significantly different between groups (no delirium/delirium) in univariate analysis.
\*significant difference between groups, p<.05
Abbreviations: APACHE=Acute Physiology and Chronic Health Evaluation, BMI=Body Mass Index (calculated with height and weight at admission), CKD=Chronic Kidney Disease includes End Stage Renal Disease, COPD=Chronic Obstructive Pulmonary Disease, ICU=Intensive Care Unit, SD=Standard Deviation

### Delirium and Coma in Critically Ill Patients with COVID-19

Delirium occurred in 73.6% (106/144) of patients in the study, whereas delirium or coma occurred in 76.4% (110/144). Forty-four percent of patients experienced coma. Of patients with delirium, 63.2% were positive on the first CAM-ICU assessment, and 36.8% developed delirium on a subsequent CAM-ICU screening. As shown in Table 1, patients with delirium had higher median APACHE-II severity of illness scores (19, IQR: 15–26 vs. 13.5, IQR: 9–17, p< 0.001) and were more likely to be mechanically ventilated (91.5% vs. 21.1%, p< 0.001). Patients with delirium had lower PaO_2_:FiO_2_ ratios (90, IQR:70.0–140.0 vs. 134.4, IQR: 95.0–188.9, p = 0.016) and lower Glasgow Coma Scale scores (10.0, IQR: 6.0–14.0 vs. 15.0, IQR: 14–15.0, p< 0.001) during the first 24 hours of ICU admission compared to those without delirium. Patients with delirium also had decreased level of consciousness by RASS score (−1.6, IQR:-2.5, –0.8 vs. 0, IQR:0–0) compared to patients who did not develop delirium assessed over 14 days of ICU stay.

### Delirium Duration, Subtypes of Delirium and Delirium Severity

The median duration of delirium and coma was 7 days (IQR: 3–10) (see Table 2), and median delirium duration was 5 days (IQR: 2–7). Patients had a median RASS of –2 (IQR: –3,0) at the time of ICU admission indicating light sedation. Figure 1 shows the daily rates of patient’s delirium, coma or delirium/coma-free status for up to 14 days of ICU admission. In our study cohort, hypoactive delirium occurred in 86.8% of patients on the first CAM-ICU assessment, and the median duration of hypoactive delirium was 4 days (IQR: 2–4). Details of the subtypes of delirium are shown in Table 2 and Figure 2. In the subset of patients with delirium severity assessments (n = 73), the median CAM-ICU-7 score was 6 (IQR: 4–7) representing severe delirium.

**Table 2.** Clinical outcomes in Critically Ill Patients Admitted with COVID19 Who Developed Delirium.

| Outcome Measures |  | Patients with Delirium<br>(n=106) |
| --- | --- | --- |
| **Duration of Delirium and Coma, median (IQR)** |  |  |
|  | Delirium/coma days | 7 (3-10) |
|  | Delirium duration in days | 5 (2-7) |
|  | Coma duration in days | 1 (0-4) |
| **Subtypes of Delirium, n (%)** |  |  |
|  | Hypoactive delirium at 1<sup>st</sup> ICU assessment | 92 (86.8) |
|  | Hyperactive delirium at 1<sup>st</sup> ICU assessment | 14 (13.2) |
| **Duration of Subtypes of Delirium, median days (IQR)** |  |  |
|  | Hypoactive delirium duration days | 4 (2-6) |
|  | Hyperactive delirium duration days | 0 (0-1) |
| **Delirium Severity, median (IQR)** |  |  |
|  | CAM-ICU-7 score | 6 (4-7) |
| Delirium was defined as a positive CAM-ICU assessment in the patient medical record for up to 14 days during their ICU COVID-19 stay. Coma was defined by Richmond Agitation Sedation Score of -4 or -5. Duration of delirium was defined as number of days patient was CAM-ICU positive on either morning or afternoon assessment for up to 14 days while admitted to the ICU. Duration of coma was defined as number of days patient had coma by RASS score on either morning or afternoon assessment for up to 14 days of ICU stay. Hypoactive delirium was defined by (RASS) of -1 to -3 with positive CAM-ICU, Hyperactive delirium was defined by a RASS score of +1 to +3 with positive CAM-ICU. Delirium severity was measured using the CAM-ICU-7 in 73 patients (0-7, 0-2: no delirium; 3-5: mild to moderate delirium; 6-7: severe delirium).<br>Abbreviations: CAM-ICU and CAM-ICU-7=Confusion Assessment Method-Intensive Care Unit, ICU=Intensive Care Unit, IQR=Interquartile Range, RASS= Richmond Agitation and Sedation Scale |  |  |

**Figure 1.**
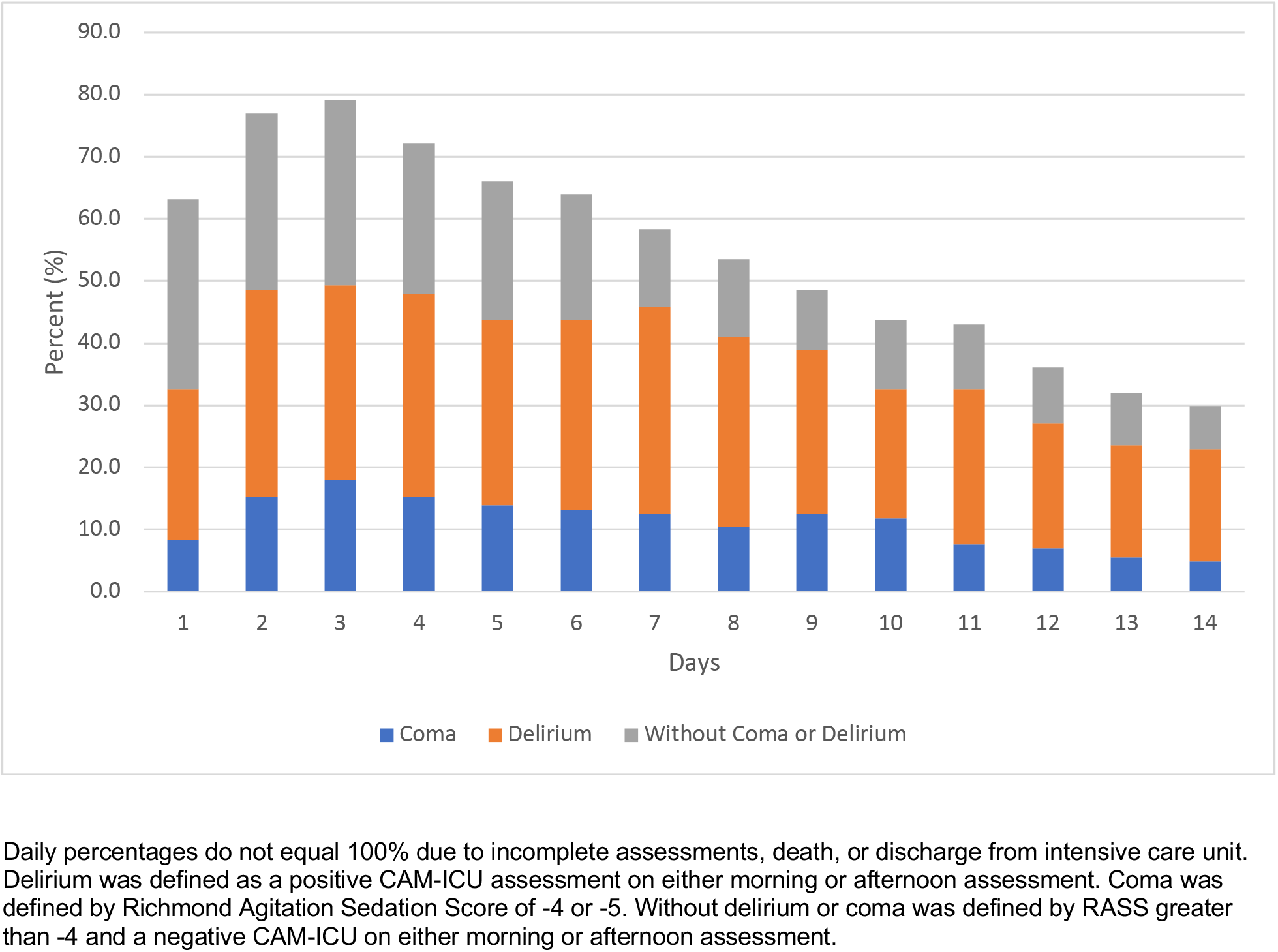
Daily Rates of Delirium, Coma, or Without Delirium/coma Status as Assessed up to First 14 Days of Intensive Care Unit Stay (n=144)

**Figure 2.**
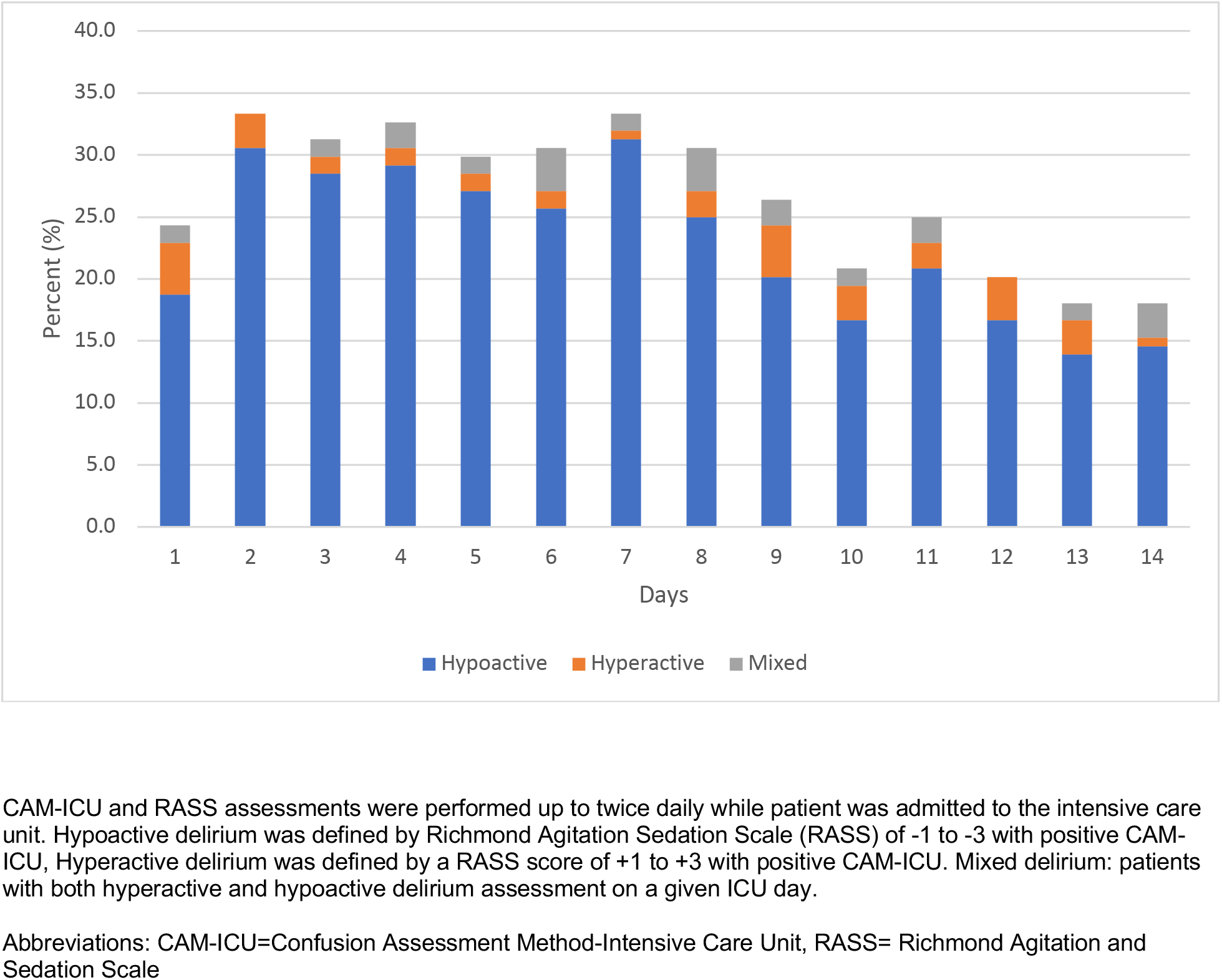
Subtypes of Delirium in Critically Ill Patients with COVID-19 Who Developed Delirium (n=106)

### Factors Associated with Delirium

Patients with delirium had greater mechanical ventilation days (median 8.7 days, IQR: 4.5–12.9 vs. 0, IQR: 0–0, p< 0.001) and ICU days (median 11.0, IQR: 6.9–15.2 vs. 3.6, IQR: 1.7-6.0, p< 0.001) compared to patients without delirium (Table 3). We did not find a significant difference in hospital mortality between COVID-19 patients with delirium and those without(26.4% vs. 15.8%, p = 0.27), as shown in Table 3. Daily rates of patient’s discharge from ICU and death over the first 14 days are shown in Supplementary Figure 2. In the logistic regression model consisting of age, receipt of mechanical ventilation, and APACHE-II scores, only mechanical ventilation was significantly associated with greater odds of developing delirium (OR: 42.1, 95% CI: 13.0–137.1, p< 0.001).

**Table 3.** Clinical outcomes of patients with COVID-19 in the Intensive Care Unit.

| **Clinical Outcomes** | **Total**<br>**(n=144)** | **Delirium**<br>**Positive**<br>**(n=106)** | **Delirium**<br>**Negative**<br>**(n=38)** | **p-value** |
| --- | --- | --- | --- | --- |
| **Median (IQR), Days** |  |  |  |  |
| Mechanical ventilation | 6.2 (0-11.3) | 8.7 (4.5-12.9) | 0 (0-0) | <.001 |
| ICU LOS | 9.7 (4.7-14.7) | 11.0 (6.9-15.2) | 3.6 (1.7-6.0) | <.001 |
| Hospital LOS | 30.5 (23-34) | 31 (25-35) | 26.5 (20-32) | .020 |
| **Mortality, n (%)** |  |  |  |  |
| In-hospital | 34 (23.6) | 28 (26.4) | 6 (15.8) | .265 |
| Table 3 describes the clinical outcomes in COVID-19 patients admitted to the ICU. The overall cohort is described then divided by delirium status (positive CAM-ICU). Univariate testing was completed to investigate statistical significance. |  |  |  |  |
| Abbreviations: ICU=Intensive Care Unit, IQR=Interquartile Range, LOS=Length of Stay |  |  |  |  |

## Discussion

In this observational study of COVID-19 patients admitted to the ICU at 2 large hospitals, 74% of patients experienced delirium, delirium occurred early in the ICU course (within the first two days), and the abnormal cognitive states of delirium or coma persisted for median length of one week. In addition, patients with COVID-19 experienced severe delirium, and invasive mechanical ventilation was associated with a marked increase in odds of delirium. While mortality rates did not statistically differ by delirium status likely due to the small sample size of patients without delirium, we found mortality to be 10% higher in patients with delirium. To the best of our knowledge, our study is the first to describe delirium rates, duration and severity in critically ill patients with COVID-19 using standardized delirium assessment tools. Due to the increased risk of mortality and morbidity following delirium, including the development of long-term cognitive impairment and post intensive care syndrome, this study has important implications for clinical practice, the recovery of patients with COVID-19 admitted to intensive care, public health decision making, and even future research priorities.^26,27^

Our study findings represent a significant departure from recently reported trends in rates of ICU delirium, including rates of mechanical ventilation (36%), delirium (22.7%), and coma (24.0%) at our own center during the influenza pandemic occurring in 2009–2010 (see Supplementary Table 2). Reductions in the prevalence of ICU delirium from a historical high of 80% to rates of 16.5–33% have been reported over the past two years.^13,16,17,19^ These reductions were likely linked to the implementation of the Society of Critical Care Medicine’s PADIS guidelines, multi-disciplinary bundled protocols for delirium prevention (ABCDEF), avoidance of deliriogenic agents, increasing use of non-invasive ventilation strategies and heated high-flow nasal cannula devices leading to reduced rates of invasive ventilation, and increasing clinician awareness regarding the harms of delirium.^17,28–30^ The COVID-19 crisis has seriously challenged these care improvements as pre-established multi-disciplinary care models of ICU care become disrupted and health systems are overwhelmed with critically ill patients requiring invasive mechanical ventilation. In addition, it has been proposed that COVID-19 may directly infiltrate the central nervous system leading to delirium.^31–33^ A recent meta-analysis on neuropsychiatric symptoms associated with severe coronavirus infections concluded that signs of delirium (confusion 27.9% 95% CI: 20.5–36.0, impaired concentration or attention 38.2% 95% CI: 29.0–47.9, and altered consciousness 20.7% 95% CI: 12.6–30.3) were common in Severe Acute Respiratory Syndrome (SARS) and Middle East Respiratory Syndrome (MERS).^34^ Further, as recent studies have reported neurologic symptoms such as anosmia in COVID-19 and SARSCoV-2 has been identified in cerebrospinal fluid as well as brain tissue, the neurotoxic impact of COVID-19 is increasingly plausible.^31–33,35^ The possible pathways for neuronal damage due to COVID-19 require additional study.

While effective pharmacological therapies for treatment of COVID-19 as well as delirium are not yet available, our study sheds light on an alarming burden of delirium and coma in patients admitted to the ICU and the need for continued efforts on delirium prevention. Following and implementing evidence-based ICU practices (such as the ABCDEF bundle) to minimize delirium occurrence and severity under the pandemic conditions will likely remain an ongoing challenge.^30^ The continued use of screening tools for delirium and delirium severity can also provide bedside clinicians with dynamic assessments to measure the impact of interventions in real-time.^9,12^ As resources shrink in the face of the pandemic and the health care response disrupts, it is imperative to continue to follow and implement time-tested evidence-based practices. Finally, delirium in critically ill patients has been associated with long-term cognitive decline.^10,36^ If other studies confirm higher rates of delirium in COVID-19 ICU patients, longitudinal follow-up will be crucial to understand the full impact of COVID-19 and understand the pathophysiology of COVID-19 related delirium.

Our study does have important limitations. This analysis is limited by its reliance on data from the medical record including clinician-administered delirium assessments. The limitation of clinician-administered delirium assessments has been minimized by the rigorous implementation and continued education on the CAM-ICU and CAM-ICU-7 at the participating institutions. Our analysis also does not include medication exposure data, adherence to the ABCDEF bundle at the patient level, education levels, baseline functional status, or baseline cognitive function and therefore we are unable to fully explain the rates of delirium seen in our study. Our analysis is also limited to delirium and coma assessments performed in the first fourteen days of ICU stay, and therefore we are unable to describe the trajectory of delirium and coma for the duration of the hospitalization in this report. Strengths of the study include incorporation of delirium severity data, a racially and socioeconomically diverse cohort of patients and protocolized delirium assessments conducted by bedside clinicians at two high volume and high acuity centers.

## Conclusion

We found that in contrast to recent rates of delirium in ICU patients, 74% of patients with COVID-19 develop delirium which persists for approximately 1 week, and occurs at high severity. Invasive mechanical ventilation is significantly associated with delirium development. Given these findings, continued attention to prevent and manage delirium is critical.

## Data Availability

A de-identified dataset may be available from the corresponding author on reasonable request.

## Notes

### Competing Interest Statement

The authors have declared no competing interest.

### Funding Statement

Babar Khan, Sujuan Gao, and Anthony Perkins are supported through NIA R01 AG 055391, R01 AG 052493 and NHLBI R01 HL131730. Anthony Perkins is also supported by NIA grants 1K23AG062555-01 and R01AG056325. Roberto Machado is supported by 1R01HL111656, 1R01HL127342 and 1R01HL133951. Sophia Wang is supported by K23AG062555-01. Edward Marcantonio is supported by grants R01AG044518 and K24AG035075 from the NIA. Malaz Boustani received funding from NIA R01AG034205 and disclosed that he has ownership equity in two for profit companies, Preferred Population Health Management and RestUp. The products and services of the two companies are not related to the research activities of the paper.

### Author Declarations

The study received ethical approval from the Institutional Review Board at Indiana University.

